# Identification of a Novel *Plasmodium falciparum Kelch13* A676T Variant and high Chloroquine and Sulfadoxine-Pyrimethamine Resistance in Cibitoke, Burundi

**DOI:** 10.1101/2025.06.22.25330092

**Authors:** Tolulope Kayode, David Niyukuri, Aurel Holzschuh, Gustavo Da Silva, Tiffany Huwe, Anita Lerch, Joseph Nyandwi, Cristian Koepfli

## Abstract

Antimalarial resistance, including reduced susceptibility to artemisinin-based combination therapies, is increasing in Africa. 157 *P. falciparum* isolates from Cibitoke, Burundi, were sequenced for ten resistance markers (including background loci associated with *kelch13*-mediated artemisinin partial resistance) using Nanopore sequencing. *Plasmepsin*-2 and *pfmdr1* copy number variations were assessed by digital PCR. No validated *pfkelch13* mutations linked to artemisinin partial resistance were detected. A novel *pfkelch13* mutation, A676T, was identified in one isolate, adjacent to the validated A675V mutation reported in neighboring East African countries. Mutant alleles were observed at several background loci previously implicated in artemisinin resistance in Southeast Asia, including *pffd* D193Y (7.0%), *pfmdr2* I492V (31.8%), *pfatg18* T38I (3.2%), and *pfarps10* V127M/D128H (2.6%), while *pfpx1* (C1484F) and *pfmdr2* (T484I) remained wild type. However, their functional role in Africa remains uncertain. Sulfadoxine-pyrimethamine (SP) resistance was widespread. In single-clone infections, the quintuple *pfdhfr/pfdhps* haplotype predominated (59.6%). Chloroquine resistance–associated *pfcrt* IET triple mutant haplotypes at codons 74-76 were detected in 94.3% of isolates. *pfmdr1* haplotypes defined by codons 86, 184, and 1246 were dominated by NYD (56.3%) and NFD (40.4%). No *pfpm2* duplications were observed, while *pfmdr1* copy number amplification was rare (1.8%). These findings reveal a high prevalence of mutations associated with SP resistance, posing a threat to the efficacy of intermittent preventive treatment in pregnancy and seasonal malaria chemoprevention in Burundi. The identification of the *pfkelch13* A676T variant highlights the importance of continued genomic surveillance. However, the role of this mutation in artemisinin resistance remains to be determined.

## INTRODUCTION

Malaria remains a major global health challenge, with an estimated 282 million cases and 610,000 deaths reported in 2024[1]. Sub-Saharan Africa bears the highest burden, accounting for 94% of malaria cases and 95% of deaths, with children under five years disproportionately affected[2]. In addition to treatment of clinical cases, antimalarial drugs are central for prevention. The emergence and spread of drug-resistant *Plasmodium falciparum* is a major threat to malaria control and elimination[3].

Artemisinin-based combination therapies (ACTs), pairing fast-acting artemisinin derivatives with longer-acting partner drugs to ensure parasite clearance,[4,5]. Sulfadoxine–pyrimethamine (SP) remains the standard for intermittent preventive treatment in pregnancy (IPTp) and for seasonal malaria chemoprevention (SMC) in children[6].

Mutations in *pfkelch13* are the primary markers of artemisinin partial resistance (ART-R). Yet, not all mutations in *pfkelch13* contribute to resistance, and delayed clearance has occasionally been observed in parasites lacking these mutations[7]. The functional significance of candidate *pfkelch13* mutations can be assessed by tracking parasite clearance in treated patients [8], through *in vitro* ring-stage survival assays (RSA) [7] or by generating isogenic parasite lines using CRISPR/Cas9 genome editing and evaluating their susceptibility with RSA [9,10]. Validated *pfkelch13* mutations associated with artemisinin partial resistance (ART-R) in Southeast Asia include key propeller domain variants such as C580Y, R539T, Y493H, and I543T, among others [11]. In Africa, several *kelch*13 variants have been reported, including the validated resistance mutation R561H, C469Y, P553L, R622I and A675V [12–19]. R561H has been observed in Rwanda[12,20], Tanzania[14,15] and Eritrea[21]. Additionally, C469Y and A675V, both associated with delayed parasite clearance, have been detected in Uganda [16].

Background mutations in genes such as *pfarps10*, *pffd*, *pfcrt*, *pfap2mu*, *pfatg18*, *pfpx1*, and *pfmdr2*, are thought to facilitate the emergence of *pfkelch13*-mediated resistance, particularly in Southeast Asia [22,23]. While some of these mutations have been reported in Africa [12,24–27], their contribution to resistance in African parasites remains unclear. Beyond *pfkelch13*, mutations in *pfcoronin* have been reported as a potential marker of partial artemisinin resistance [28], although their role is not yet fully established.

Resistance to ACT partner drugs further complicates treatment outcomes. Mutations in *pfcrt* are associated with chloroquine resistance and have also been linked to reduced susceptibility to piperaquine, while polymorphisms in *pfmdr1* and *pfcrt* are candidate markers amodiaquine resistance [29–32]. Amplification of the *plasmepsin 2/3* gene cluster has been linked to piperaquine resistance in Southeast Asia [33]. In contrast, *pfmdr1* copy number variation has been explored as a potential marker of altered responses to lumefantrine, although it is not considered a validated marker of resistance [34].

Sulfadoxine–pyrimethamine (SP) remains widely used across sub-Saharan Africa for preventive interventions, including intermittent preventive treatment in pregnancy (IPTp) and seasonal malaria chemoprevention (SMC). However, resistance to SP is driven by the spread of quintuple and sextuple *pfdhfr/pfdhps* haplotypes, which are approaching fixation in several East African countries, including Tanzania, Kenya, and Uganda [35–39].

In Burundi, nearly 80% of the population remains at risk, and malaria cases rose from 6.8 million in 2021 to over 8.2 million in 2022[40]. Burundi’s malaria control strategy relies on SP for IPTp, and ACTs, primarily artemether–lumefantrine (AL), for the treatment of uncomplicated malaria [40,41]. Given its proximity to regions with confirmed ART-R, including Rwanda[12], Uganda[16], the Democratic Republic of Congo (DRC) [42] and Tanzania [14], Burundi faces heightened risk of resistant parasite introduction and local establishment. However, data on *pfkelch13* polymorphisms and other molecular resistance markers in Burundi remains scarce.

This study employed multiplexed PCR-based targeted amplicon sequencing to characterize single nucleotide polymorphisms (SNPs) in key antimalarial drug resistance markers, alongside digital PCR (dPCR) to assess copy number variation (CNV) in *Plasmodium falciparum* isolates from Burundi. We identified a novel *pfkelch13* mutation and a high prevalence of sulfadoxine–pyrimethamine and chloroquine resistance–associated variants. This study provides new insights into the molecular epidemiology of antimalarial resistance markers in *Plasmodium falciparum* from Burundi.

## MATERIALS AND METHODS

### Ethical approval

This study was conducted in compliance with ethical standards and received approval from the Comité National d’Ethique pour la protection des êtres humains sujets de la recherche biomédicale et comportementale of Burundi (approval reference: CNE/03/2021) and the Institutional Review Board of the University of Notre Dame (approval reference: 21-02-6446). All adult participants gave informed written consent prior to sample collection, and minors’ legal guardians provided consent in writing.

### Study Area, sample collection, and molecular screening

Burundi experiences high, year-round transmission of *P. falciparum* malaria, with moderate seasonal fluctuations. Samples for this study were collected during the dry season in June 2021 from Cibitoke Province, located in northwestern Burundi, bordering the Democratic Republic of the Congo to the west and Rwanda to the north. Notably, *pfkelch13* mutations associated with partial artemisinin resistance have been reported in parts of these neighboring countries. The sample collection was part of a previous evaluation of rapid diagnostic tests (RDT) in both clinical and subclinical populations [43]. Briefly, clinical samples were obtained from individuals presenting to healthcare facilities with suspected malaria, while subclinical samples were collected from community members using a cross-sectional, convenience-based sampling approach. Approximately 200 μL of blood was obtained via finger prick into EDTA tubes and stored at -20°C until molecular analyses were conducted. Molecular screening for *P. falciparum* was conducted using the *var*ATS qPCR assay, as described previously [43].

### Multiplexed PCR for marker amplification

Multiplexed PCR was employed to amplify defined genomic regions of *Plasmodium falciparum* genes associated with antimalarial drug resistance (Table 1), followed by amplicon sequencing. NF54, Dd2, HB3, and KH004 reference strains were included as positive controls in the multiplex PCR. A comprehensive list of primer sequences is provided in Supplementary Table S1.

**Table 1.** Target Genes, Amplicon Sizes, and Codons Included in the Multiplex PCR Panel for Antimalarial Drug Resistance.

| Gene | Amplicon size (bp) | Target Codon(s)/Region | Relevance |
| --- | --- | --- | --- |
| *arps10* | 426 | V127M, D128H | ART-R background mutations |
| *px1* | 208 | C1484F | ART-R background mutations |
| *fd* | 262 | D193Y | ART-R background mutations |
| *atg18* | 222 | T38I | ART-R background mutations |
| *mdr2* | 216 | T484I, I492V | Putative partner drug response marker |
| *mdr1* | 78 | N86Y | Partner drug response modulation |
| *mdr1* | 134 | Y184F | Partner drug response modulation |
| *mdr1* | 227 | S1034T/C, N1042D | Partner drug response modulation |
| *mdr1* | 177 | F1226Y, D1246Y | Partner drug response modulation |
| *dhps* | 92 | S436A/F, A437G | Sulfadoxine resistance |
| *dhps* | 289 | K540E, A581G, A613T/S | Sulfadoxine resistance |
| *dhfr* | 269 | N51I, C59R, S108N | Pyrimethamine resistance |
| *dhfr* | 255 | I164L | Pyrimethamine resistance |
| *crt* | 311 | M74I, N75E, K76T, T93S, H97L | Chloroquine resistance |
| *crt* | 339 | M343L, C350R, G353V, I356T | Chloroquine resistance |
| *kelch13* | 218 | Codons 435-506 | ART-R |
| *kelch13* | 285 | Codons 520-580 | ART-R |
| *kelch13* | 310 | Codons 600-699 | ART-R |

ART-R: Artemisinin resistance

To amplify markers, three separate PCR reactions were performed using distinct primer mixes: PM 1, PM 2, and PM 3 (Supplementary Table S2). All primers were synthesized by Sigma-Aldrich (St. Louis, MO, USA). Each reaction was prepared in a total volume of 25 μL, consisting of 14.25 μL of PCR-grade water, 5 μL of 5X KAPA HiFi Buffer (1X final concentration), 0.75 μL of 10 mM KAPA dNTP Mix (0.3 mM final concentration), 0.5 μL of a primer mix (PM 1, PM 2, or PM 3), 0.5 μL of HiFi HotStart DNA Polymerase (1 U/μL, final concentration 0.5 U; Roche, Basel, Switzerland), and 4 μL of DNA template. PCR amplifications were conducted on a BioRad T100 Thermal Cycler (Bio-Rad Laboratories, Hercules, CA, USA) and consisted of an initial denaturation at 95°C for 3 minutes, followed by 35 cycles of denaturation at 98°C for 15 seconds, annealing at primer-specific temperatures (58°C for PM 1, 62°C for PM 2, and 51°C for PM 3) for 15 seconds, extension at 72°C for 30 seconds, and a final extension at 72°C for 2 minutes.

### Library Preparation and Sequencing

Amplicons from the three PCR reactions were pooled in equimolar concentrations for downstream sequencing preparation. Library preparation was conducted using the Oxford Nanopore Technologies (ONT) SQK-NBD114.24 and 96 kit, following the manufacturer’s protocol (version NBA_9170_v114_revL_15Sep2022) with modifications as described earlier [44]. Briefly, for the end-prep step, incubation times were extended to 15 minutes each at 20°C and 65°C. End-prepped DNA was purified using a 1.8X bead-to-sample ratio of AMPure XP beads (Beckman Coulter, Brea, CA, USA) and resuspended in 15 μL of nuclease-free water. 3.75 μL of end-prepped DNA was used instead of the standard 0.75 μL for the native barcoding ligation, and the incubation time was extended to 30 minutes from 20 minutes. Barcoded samples were purified using a 1.4X bead-to-sample ratio of AMPure XP beads, replacing the standard 0.4X ratio. During the adapter ligation and clean-up step, the incubation time was increased to 30 minutes, and pooled barcoded samples were purified using 0.8X AMPure XP beads. Short fragment buffer was used for all wash steps during purification.

The final pooled library was quantified using a Qubit fluorometer (Thermo Fisher Scientific, Waltham, MA, USA) and diluted in ONT elution buffer to approximately 130 fmol before being loaded onto R10.4.1 flow cells. Sequencing was conducted on a MinION Mk1C instrument (ONT) using MinKNOW software (distribution version 23.07.12, core version 5.7.5, and configuration version 5.7.11).

### Bioinformatics Analysis of Sequencing Data

Nanopore sequencing data (*.pod5 files) were basecalled using Dorado (v0.8.3; https://github.com/nanoporetech/dorado) with the super accuracy model dna_r10.4.1_e8.2_400bps_sup@v5.0.0. Previously established thresholds and flags were used[44,45]. Briefly, the --no-trim flag was applied to retain full-length reads. To ensure high data quality, a minimum Q-score threshold of 20 (--min-qscore 20), corresponding to an accuracy of ≥99%, was applied to filter reads for downstream analysis. Quality-filtered reads were demultiplexed using Dorado with double-ended demultiplexing enabled (--barcode-both-ends) to minimize false-positive assignments. A summary of the basecalling results, including reads below the Q-score threshold, was generated using the Dorado summary command, and quality metrics were further assessed with NanoStat.

Nanopore sequencing data were processed as described earlier [44,45]. In short, haplotypes were inferred from the resulting FASTQ files using the R packages HaplotypR (v0.5.0; https://github.com/lerch-a/HaplotypR) and DADA2 (v1.26.0) [46,47]. Reads were demultiplexed by marker using HaplotypR’s demultiplexByMarkerMinION() function, and sequences with ambiguous bases or incorrect lengths were excluded.

Haplotypes were inferred using DADA2’s learnError() and dada() functions, with sequence abundance tables generated using makeSequenceTable(). To ensure the integrity of the data, chimeric haplotypes were identified and removed using removeBimeraDenovo(). Additional filtering excluded sequences with singletons or ambiguous bases. For each sample, markers were included if they had a minimum coverage of 10 reads. Haplotype inference required at least 50 supporting reads per haplotype and a within-host frequency of ≥1%

Haplotypes were analyzed via visual inspection and alignment against annotated *P. falciparum* 3D7 reference genomes using Geneious Prime (v2025.0.3). Reference sequences used were *pfkelch13* (PF3D7_1343700), *pfdhfr* (PF3D7_0417200), *pfmdr1* (PF3D7_0523000), *pfcrt* (PF3D7_0709000), *pfpx1* (PF3D7_0720700.1), *pfdhps* (PF3D7_0810800), *pfatg18* (PF3D7_1012900), *pffd* (PF3D7_1318100), *pfmdr2* (PF3D7_1447900), and *pfarps10* (PF3D7_1460900).

Haplotypes that failed to align to their respective reference sequences were excluded from analysis. In several targets (*pfarps10, pfatg18, pffd*, and *pfcrt* codons 343-356), insertions or deletions (indels) were observed within the amplified regions, mostly downstream of the focal resistance-associated codons. These indels did not affect SNP-based resistance classification but contributed to overall haplotype diversity (Supplementary Figures S1-S4).

For validation, samples in which *pfkelch*13 SNPs were detected were re-amplified in supplicate using the same primer sets targeting three overlapping regions but amplified independently without the other gene targets. Amplicons were sequenced on a MinION Mk1C platform. In addition, a *pfkelch1*3-specific simplex PCR was performed using the outer primer pair flanking codons 400–699, thereby generating a single contiguous amplicon spanning the entire region. This reaction was conducted in duplicate, and the resulting PCR products were sequenced separately via Sanger sequencing. Variants were considered verified only if identical SNPs were consistently detected across sequencing runs.

### Digital PCR Analysis of Gene Copy Number Variations in *pfmdr1* and *pfpm2*

Digital PCR assays were developed to quantify *pfpm2* and *pfmdr1* and β*-tubulin* as single-copy control gene. Primer and probe sequences are listed in Supplementary Table S4. Each reaction was prepared in a total volume of 12 μL, comprising 3 μL of QIA-PCR Master Mix (1X final concentration), 1.08 μL each of 10 μM forward and reverse primers, 0.3 μL of 10 μM probes, 1 μL of restriction enzyme, and 2 μL of DNA template, and 1.08 μL of nuclease-free water. For the *pfpm2* assay, HinfI (10 U) was used, while AluI (10,000 U/mL) was included for the *pfmdr1* assay. Assays were run on the QIAcuity One (Qiagen, Germany). Cycling conditions for both assays included an initial denaturation at 95°C for 2 minutes, followed by 49 cycles of 95°C for 15 seconds and 60°C for 1 minute. Copy number variation (CNV) was calculated as the ratio of the parasite density of the target gene to that of the β*-tubulin* control gene. A CNV ratio of ≥1.5 was considered a gene duplication.

## RESULTS

### Reads Distribution and Sequencing performance

Sequencing read depth (log10-transformed) across all targets is shown in Fig. 1, demonstrating generally robust amplification with variable coverage across markers. Supplementary Table S5 summarizes genotype calls for each marker, including wild-type, mutant, and mixed infections. Most samples exceeded the minimum threshold of 10 reads per marker, with 86–100% of targets achieving at least 50 reads.

**Figure 1.**
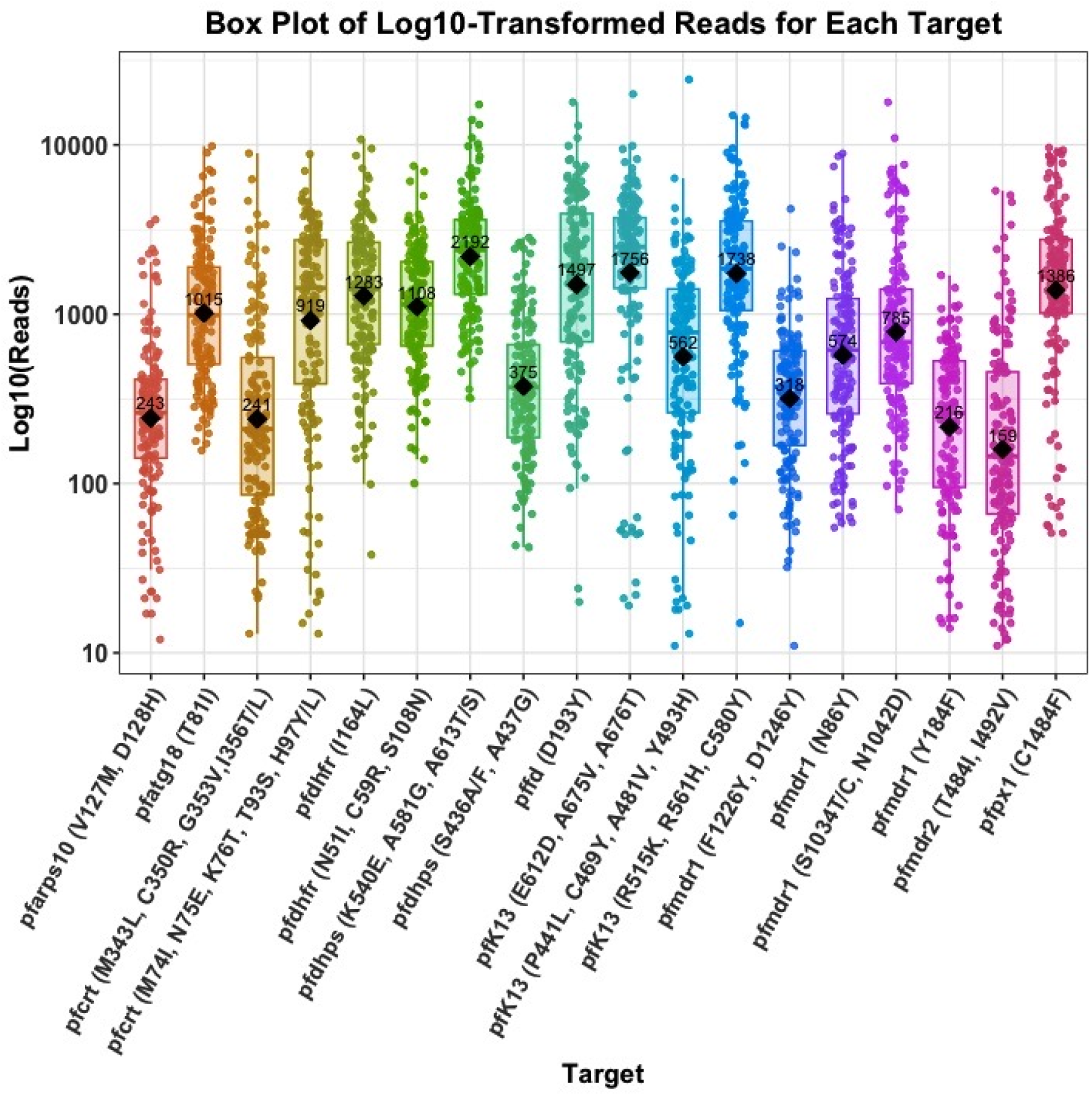
Read coverage and distribution across genetic markers. Log10-transformed read counts for each amplified genetic marker. Boxplots show the distribution of reads per target across samples.

To assess the relationship between parasite density and sequencing performance, Spearman’s rank correlation analysis was performed. No significant correlation was observed between parasite density and total sequencing depth across all markers (ρ = −0.03, p = 0.70), indicating that overall sequencing performance was not strongly influenced by parasite density. Consistent with this, read counts were observed across the full range of parasite densities, including low-density infections, although variability in read depth was evident.

At the level of individual amplicons, correlations between parasite density and read depth varied by marker. Moderate positive correlations were observed for a subset of loci, including several pfk13, pfcrt, and pfmdr1 targets, whereas other markers showed weak, negligible, or negative correlations. These findings indicate locus-specific variability in amplification efficiency rather than a consistent density-dependent effect on sequencing depth. These relationships are shown in Supplementary Fig. S5.

### Polymorphisms in *pfkelch13*

All 157 *P. falciparum* samples were successfully genotyped for *pfkelch*13 mutations, identifying one non-synonymous mutation, A676T, and two synonymous mutations (S477S and T685T) (Supplementary Figures S6 and S7). Mutations were confirmed by Sanger sequencing. The A676T (GCC→ACC) mutation has not been previously reported. A different substitution at the same codon (A676D) has been described in parasites from China-Myanmar border [48–53]. The A676T variant is located adjacent to A675V (GCC→GTC), a WHO-validated *pfkelch*13 mutation associated with partial artemisinin resistance and documented in East African parasite populations, including Uganda, Rwanda and Kenya [16,54–56]. However, there is currently no functional, clinical, or *in vitro* evidence linking A676T to artemisinin resistance, and its significance remains to be determined.

### Polymorphisms in artemisinin-predisposing background loci

No mutations were detected at *pfpx1* codon 1484 or *pfmdr2* codon 484, as all successfully genotyped isolates (157/157, 100%) carried the wild-type allele at both loci. A polymorphism was observed at *pffd* codon D193Y, where 54.1% (85/157; 95% CI 46.0-62.1) of isolates were wild type, 7.0% (11/157; 95% CI 3.5-12.2) carried the mutant allele, and 38.9% (61/157; 95% CI 31.2-46.9) exhibited mixed-allele infections. At *pfatg18* codon T38I, 88.5% (139/157; 95% CI 82.5-93.1) of isolates were wild type, 3.2% (5/157; 95% CI 1.0-7.3) carried the mutant allele, and 8.3% (13/157; 95% CI 4.5-13.7) showed mixed-allele infections.

Similarly, a polymorphism was detected at *pfarps10* codons V127M/D128H, with 88.3% (136/154; 95% CI 82.2-92.9) wild type, 2.6% (4/154; 95% CI 0.7-6.5) mutant, and 9.1% (14/154; 95% CI 5.1-14.8) mixed-allele infections. A polymorphism was observed at *pfmdr2* codon I492V, where 47.1% (74/157; 95% CI 39.1-55.2) of isolates were wild type, 31.8% (50/157; 95% CI 24.6-39.7) carried the mutant allele, and 21.0% (33/157; 95% CI 14.9-28.2) exhibited mixed-allele infections.

### Polymorphism in *pfcrt* and *pfmdr1*

All 157 *Plasmodium falciparum* samples were successfully genotyped for the *pfcrt* gene. Mutant or mixed alleles were detected at codons M74I, N75E, and K76T in 94.3% of samples (148/157; CI 89.4-97.3) (Table 2). All samples were wild type at codons T93S, M343L, C350R, and G353V, while mixed alleles were detected at H97Y/L in a single sample (0.6%). The I356T mutation was detected in 8.3% of samples (13/157; CI 4.5-13.7) comprising 12 pure mutant and one mixed-allele infection.

**Table 2.**
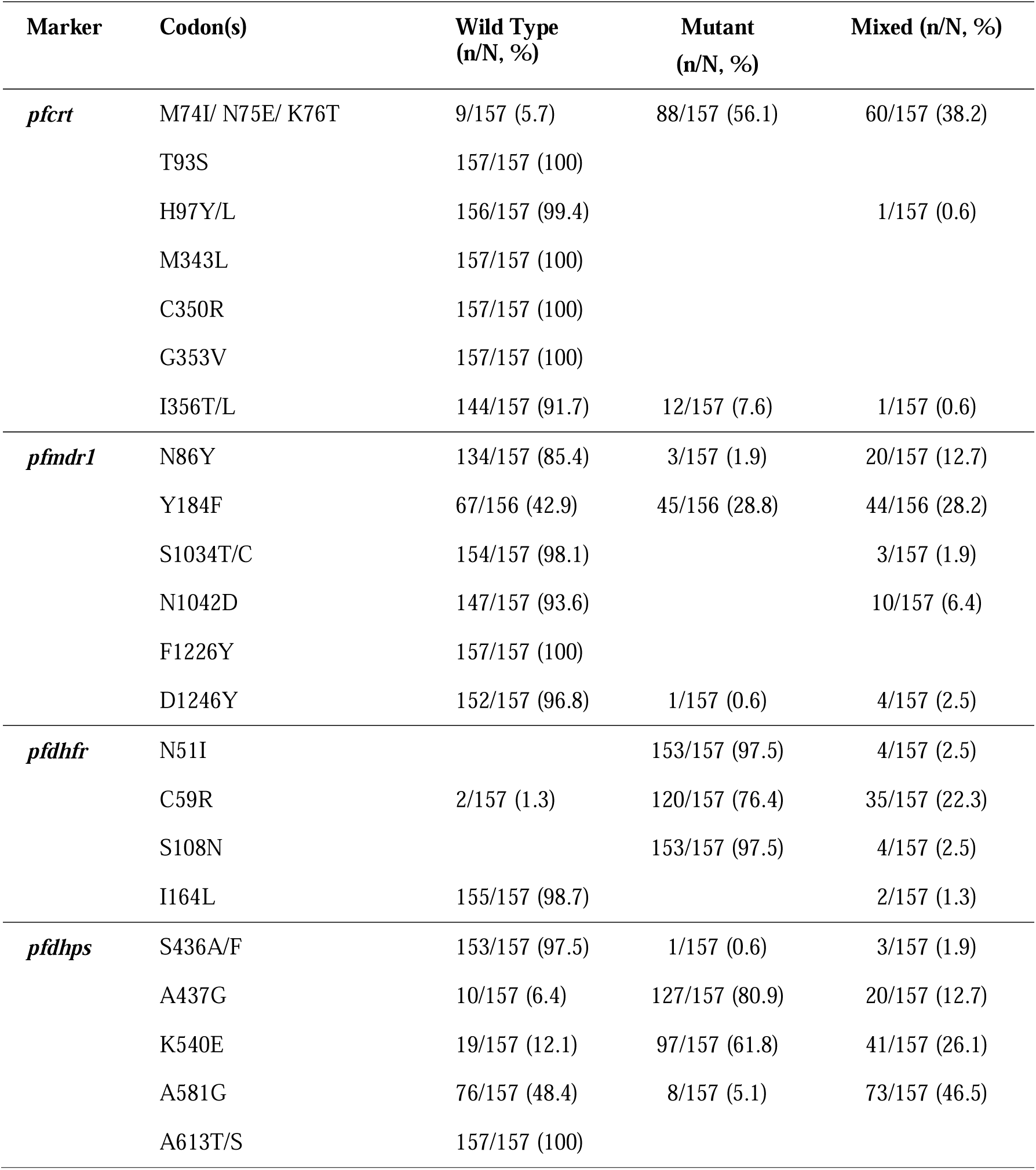
Mutations detected in *pfcrt*, *pfmdr1*, *pfdhps*, and *pfdhfr* markers.

| Marker | Codon(s) | Wild Type<br>(n/N, %) | Mutant<br>(n/N, %) | Mixed (n/N, %) |
| --- | --- | --- | --- | --- |
| *pfcr*t | M74I/ N75E/ K76T | 9/157 (5.7) | 88/157 (56.1) | 60/157 (38.2) |
|  | T93S | 157/157 (100) |  |  |
|  | H97Y/L | 156/157 (99.4) |  | 1/157 (0.6) |
|  | M343L | 157/157 (100) |  |  |
|  | C350R | 157/157 (100) |  |  |
|  | G353V | 157/157 (100) |  |  |
|  | I356T/L | 144/157 (91.7) | 12/157 (7.6) | 1/157 (0.6) |
| *pfmdr*1 | N86Y | 134/157 (85.4) | 3/157 (1.9) | 20/157 (12.7) |
|  | Y184F | 67/156 (42.9) | 45/156 (28.8) | 44/156 (28.2) |
|  | S1034T/C | 154/157 (98.1) |  | 3/157 (1.9) |
|  | N1042D | 147/157 (93.6) |  | 10/157 (6.4) |
|  | F1226Y | 157/157 (100) |  |  |
|  | D1246Y | 152/157 (96.8) | 1/157 (0.6) | 4/157 (2.5) |
| *pfdhfr* | N51I |  | 153/157 (97.5) | 4/157 (2.5) |
|  | C59R | 2/157 (1.3) | 120/157 (76.4) | 35/157 (22.3) |
|  | S108N |  | 153/157 (97.5) | 4/157 (2.5) |
|  | I164L | 155/157 (98.7) |  | 2/157 (1.3) |
| *pfdhps* | S436A/F | 153/157 (97.5) | 1/157 (0.6) | 3/157 (1.9) |
|  | A437G | 10/157 (6.4) | 127/157 (80.9) | 20/157 (12.7) |
|  | K540E | 19/157 (12.1) | 97/157 (61.8) | 41/157 (26.1) |
|  | A581G | 76/157 (48.4) | 8/157 (5.1) | 73/157 (46.5) |
|  | A613T/S | 157/157 (100) |  |  |

In the *pfmdr1* gene, mutations were observed at codons N86Y in 14.6% of samples (23/157; CI 9.5-21.2), Y184F in 57.1% (89/156; 48.9-64.9), S1034T/C in 1.9% (3/157; 0.4-5.5), N1042D in 6.4% (10/157; 3.1-11.4), and D1246Y in 3.2% (5/157; 1.0-7.3), including both pure mutant and mixed-allele infections (Table 2). All samples were wild type at codon F1226Y.

Among single-clone infections with complete *pfmdr1* haplotypes at codons 86, 184, and 1246 (n = 103), the predominant haplotype was NYD (N86 + Y184 + D1246), detected in 56.3% (58/103; 46.2-66.1) of samples, followed by NFD (N86 + F184 + D1246) in 40.4% (43/103; 32.1-51.9). Less frequent haplotypes included YYD in 1.9% (2/103; 0.2-6.8) and NFY in 1.0% (1/103; 0.0-5.3) of samples (Table 3).

**Table 3.**
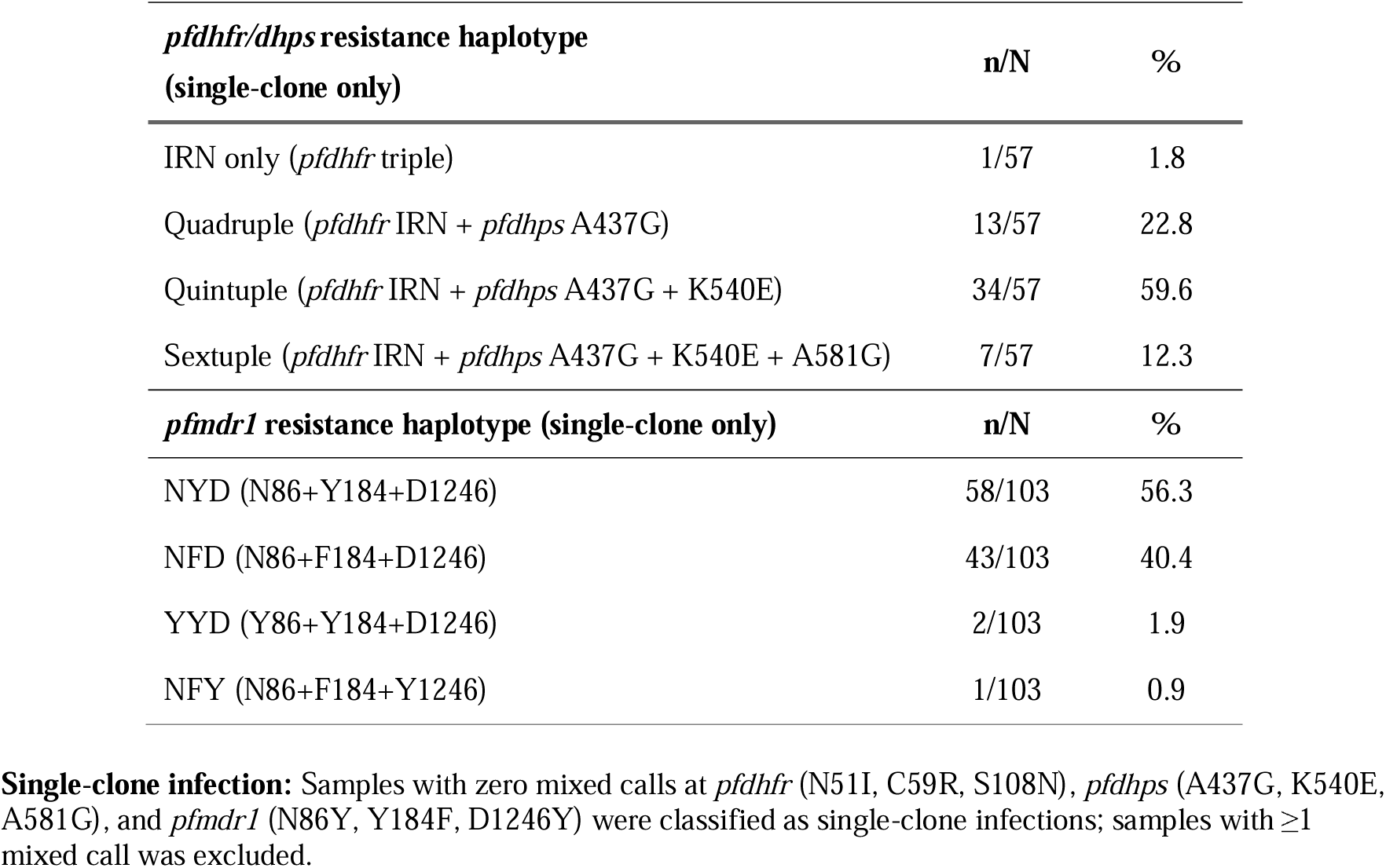
*pfdhfr/dhps* and *pfmdr1* drug resistance haplotypes in single-clone infections.

### Polymorphism in pfdhfr and pfdhps

All 157 samples were successfully genotyped for *pfdhfr*. Mutant alleles at codons N51I and S108N were detected in 100% (157/157) of isolates, including 153 pure mutant and four isolates with mixed alleles simultaneously at codons 51 and 108 (Table 2). The C59R mutation was present in 98.7% of samples (155/157; CI 95.5-99.8), comprising 120 pure mutant and 35 mixed-allele infections. The triple-mutant haplotype (IRN) was observed in 76.4% (120/157; CI 68.9–82.8) isolates. Codon I164L remained predominantly wild type, with mixed alleles detected in 1.3% (2/157; 95% CI 0.2–4.6) of samples.

For *pfdhps*, polymorphisms were observed at multiple sulfadoxine–pyrimethamine (SP) resistance–associated codons. The A437G mutation was detected in 93.6% (147/157; 95% CI 88.6–96.8) of samples, including 127 pure mutant and 20 mixed-allele infections. K540E was present in 87.9% (138/157; 95% CI 81.8–92.4) of samples, comprising 97 pure mutant and 41 mixed-allele infections. The A581G mutation was detected in 51.6% (81/157; 95% CI 43.5–59.6) of samples and occurred predominantly as mixed-allele infections. Mutations at codon S436A/F were rare (2.5% (4/157; 95% CI 0.7–6.3)), while all samples were wild type at codon A613T/S (157/157, 100.0%; 95% CI 97.7–100.0) (Table 2).

Mutations in *pfdhfr* and *pfdhps* were combined to define sulfadoxine–pyrimethamine (SP) resistance haplotypes, restricting analyses to strictly single-clone infections (no mixed calls at N51I, C59R, S108N, A437G, K540E, A581G). Among single-clone infections (n = 57), the quintuple haplotype (IRN + A437G + K540E) predominated in 59.6% (34/57; 95% CI 45.8–72.4) of samples, followed by the quadruple haplotype (IRN + A437G) in 22.8% (13/57; 95% CI 13.0–35.0). The sextuple haplotype (IRN + A437G + K540E + A581G) occurred in 12.3% (7/57; 95% CI 5.1–23.7), while IRN-only was rare (1.8% (1/57; 95% CI 0.0–9.4)) (Table 3).

### Co-occurrence of Resistance-Associated SNPs Across Antimalarial Drug Targets

UpSet analysis of SNP co-occurrence among single-clone infections revealed a dominant multidrug-resistant backbone. The most frequent intersection (n = 7) comprised the *pfdhfr* triple mutant (N51I/C59R/S108N), *pfdhps* A437G and K540E, *Pfcrt* K76T, and *mdr1* Y184F. Several related combinations were observed at lower frequencies (n = 2–3), generally retaining the *pfdhfr/dhps* resistance backbone together with *pfcrt* K76T. Additional rare intersections (n = 1) included further polymorphisms in *pfdhps, pfmdr1*, and *pfmdr2* (Fig.2).

**Figure 2.**
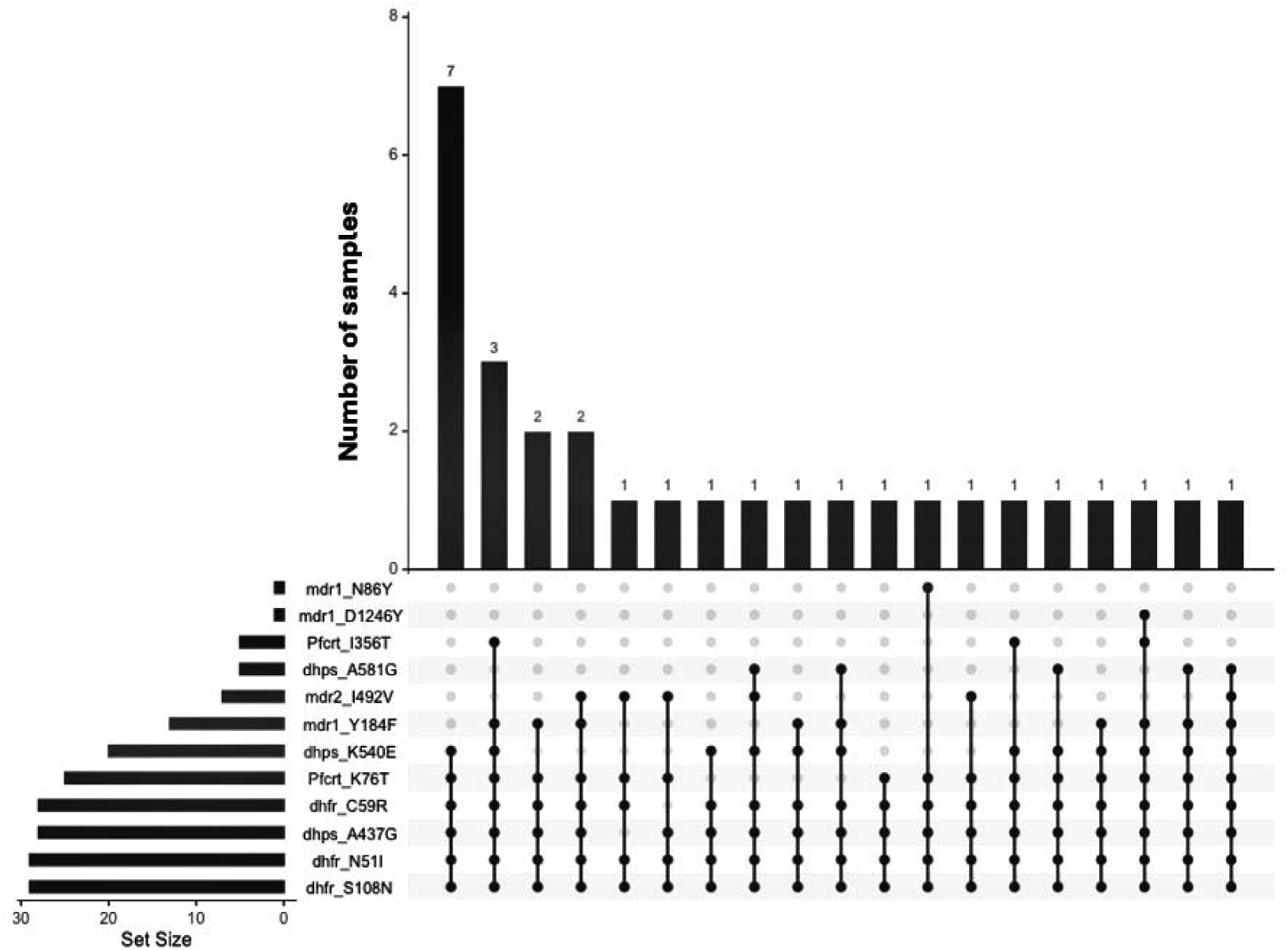
UpSet plot illustrating the co-occurrence patterns of resistance-associated single nucleotide polymorphisms (SNPs) across *pfcrt*, *pfdhfr*, *pfdhps*, *pfmdr1*, and *pfmdr2* among single-clone *Plasmodium falciparum* infections. Vertical bars represent the number of isolates sharing each specific SNP combination, while the connected black dots indicate the SNPs included in each intersection. Horizontal bars denote the overall prevalence of individual SNPs (set size). The most frequent intersection comprised the *pfdhfr* triple mutant (N51I/C59R/S108N), *pfdhps* A437G and K540E, *pfcrt* K76T, and *pfmdr1*Y184F, consistent with a multidrug-resistant genetic background. Additional lower-frequency intersections reflect further diversification through the accumulation of mutations in *pfdhps*, *pfmdr1*, and *pfmdr2*.

### Gene Duplication in *Pfmdr1* and *Pfpm2*

Copy number variations (CNVs) for *plasmepsin II (pfpm2)* and *Multidrug resistance gene 1 (pfmdr1)* were successfully assessed in all 157 samples by digital PCR. No parasites exhibited gene duplication (CNV > 1.5) for *pfpm2* (Supplementary figure 8A). In contrast, duplications were observed in *pfmdr1* in three samples (1.8%) (Supplementary figure 8B).

## DISCUSSION

This study revealed extensive chloroquine (CQ) and sulfadoxine–pyrimethamine (SP) resistance in Cibitoke Province, Burundi, alongside evidence of sustained selection on ACT partner drugs. While no validated *pfkelch13* mutations associated with artemisinin resistance were detected, the identification of a novel *pfkelch13* substitution, A676T, warrants careful evaluation.

The detection of A676T adds to evidence that the 675–676 region of the *k13* propeller domain is functionally important and tolerate amino-acid substitutions. Importantly, codon 676 lies immediately adjacent to A675V, a validated artemisinin-resistance mutation reported across multiple East African countries [20,55–59]. A different mutation at this position, A676D, has been reported in *P. falciparum* isolates from Southeast Asia, including the China–Myanmar border [48–53], where it was associated with elevated ring-stage survival and predicted intermediate parasite clearance, alongside moderate destabilization of the *K13* β-propeller domain [50,51].

However, there is currently no evidence linking the A676T variant identified in this study to artemisinin resistance and should be interpreted only as a descriptive genomic observation. Further studies, including phenotypic assays and clinical correlation analyses, are required to determine whether this mutation has any impact on parasite susceptibility to artemisinin. The identification of this novel *pfkelch13* variants underscores the importance of continued genomic surveillance to detect emerging mutations and monitor their potential epidemiological relevance.

We assessed background mutations potentially contributing to kelch13-mediated resistance. Studies from Southeast Asia have shown that mutations such as *pfarps*10 V127M, *pffd* D193Y, and *pfmdr*2 T484I occur more frequently in parasites carrying *pfkelch*13 resistance mutations and are thought to contribute to a genetic background associated with the emergence of artemisinin resistance. Two markers, *pfpx*1 (C1484F) and *pfmdr*2 (T484I), were conserved across the study population. Although polymorphisms were detected at *pffd* (D193Y), *pfatg*18 (T38I), *pfarps*10 (V127M/D128H), and *pfmdr*2 (I492V), their distribution did not resemble that reported in Southeast Asian parasites carrying *pfkelch*13 resistance mutations. These observations are consistent with previous studies [12,24,25,27,60,61] showing that some of these variants occur across African parasite populations without the resistance-associated context described in Southeast Asia. This difference might be the result of higher transmission in many parts of sub-Saharan Africa and resulting increased genetic diversity and recombination, as well as differences in historical antimalarial usage.

Patterns suggestive of selection on ACT partner drugs were observed. Artemether–lumefantrine (AL) remains the first-line treatment for uncomplicated malaria in Burundi. The high prevalence of the *pfmdr1* NFD haplotype among single-clone infections suggests sustained selection under AL pressure. This haplotype has been reported in African settings following extensive AL use [62–67], and has been associated with altered parasite responses under lumefantrine pressure [32,68–70]. However, these observations should not be interpreted as evidence of confirmed clinical resistance, and *pfmdr1* is not considered a reliable standalone marker of lumefantrine resistance, but rather a candidate gene associated with drug response.

SP remains a cornerstone of IPTp and SMC strategies in Burundi, yet the widespread prevalence of SP resistance observed in this study raises substantial concerns. The dominance of high-level resistance haplotypes, coupled with near-fixation of the most resistant *dhps* genotypes threatens the efficacy of SP-based chemoprevention. Similar resistance patterns across East Africa [71–76], contrasted with lower prevalence in much of West Africa [6,77,78], emphasize regional differences in resistance trajectories and underscore the risks posed to maternal and fetal health.

The co-occurrence of *pfdhfr* and *pfdhps* resistance mutations with *Pfcrt* K76T across most isolates indicates persistence of a combined SP and CQ resistance genetic background in this population. The frequent presence of *pfmdr1* Y184F within these combinations is consistent with ongoing drug selection pressure, including from artemether–lumefantrine. Additional low-frequency variants, including *pfdhps* 581G and *pfmdr2* I492V, further suggest ongoing diversification on this resistant background. Together, these patterns are consistent with layered drug selection, whereby historical CQ and SP pressure established a stable resistance backbone that is now being shaped by ACT partner drug selection.

Despite long-standing withdrawal of chloroquine (CQ) in Burundi, CQ resistance remains highly prevalent, as evidenced by the near fixation of the *pfcrt* IET triple mutant haplotype. Similar patterns have been reported in recent studies from northern Burundi, which also observed a high prevalence of *pfcrt* mutations associated with chloroquine resistance [79]. In several endemic countries, withdrawal of CQ has been followed by the re-emergence of CQ-sensitive *P. falciparum* parasites [80–84]. However, the continued dominance of CQ-resistant parasites in Burundi more than two decades after CQ withdrawal resembles patterns reported in parts of East Africa [85–88]. This persistence in Burundi may be partly explained by selection from amodiaquine, which was widely used in combination therapy as artesunate–amodiaquine, the national first-line treatment until 2019 [89].

This study has some limitations that should be considered when interpreting the findings. Samples were collected from a single province (Cibitoke) during a single time point (June 2021) and therefore may not capture the full spatial and temporal heterogeneity of *P. falciparum* populations across Burundi. In addition, sequencing was performed using Oxford Nanopore technology, which is associated with higher base-calling error rates compared to other next-generation sequencing platforms; however, stringent quality filtering, haplotype inference, and validation steps were applied (as described in the Methods) to mitigate this.

The novel *pfkelch13* A676T mutation identified in this study was not functionally validated, and no *in vitro* or clinical data are available to assess its potential role in artemisinin resistance. As such, these results should not be interpreted as representative of the national situation, and the significance of this variant remains to be determined. Future studies incorporating broader geographic sampling, longitudinal designs, and functional validation will be important to provide a more comprehensive understanding of antimalarial resistance dynamics in Burundi.

In conclusion, this study reveals high frequency of markers associated with chloroquine and sulfadoxine–pyrimethamine resistance and the absence of validated and candidate mutations in *pfkelch13* associated with ART-R. These findings underscore the urgency of continued surveillance, functional validation of emerging variants, and regionally coordinated responses to mitigate resistance spread and preserve the efficacy of current malaria control strategies.

## DATA AVAILABILITY

All nucleotide sequences generated in this study have been deposited in GenBank under accession numbers PV548606–PV548676. Due to GenBank length restrictions, short sequences (<50 bp) corresponding to *pfdhps* (codons 436–437) and *pfmdr1* (codons 1034–1042) are provided in Supplementary Table S6.

## ACKNOWLEDGMENTS

We thank all study participants and field teams for their contributions.

## AUTHORS’ CONTRIBUTIONS

TK, DN, JN, and CK conceived the study. DN and JN supervised sample collection and fieldwork. TK, AH, and TH conducted laboratory experiments. TK, GD, and AL performed bioinformatics analyses. TK drafted the manuscript. DN and CK secured funding, managed the project, and provided overall supervision. All authors reviewed and approved the final manuscript.

## FUNDING DEATILS

This work was supported by funds provided by the University of Notre Dame. C.K. was supported by the Bill & Melinda Gates Foundation (Grant INV-005898). T.K. received support from the Indiana Clinical and Translational Sciences Institute, funded in part by Grant Number UM1TR004402 from the National Institutes of Health, National Center for Advancing Translational Sciences, through the Clinical and Translational Sciences Award. T.K. is also supported by a postdoctoral fellowship from the Environmental Change Initiative at the University of Notre Dame, which provides partial salary support.

