## Supplementary Figures for "Identification of a Novel *Plasmodium falciparum Kelch13* A676T Variant and high Chloroquine and Sulfadoxine-Pyrimethamine Resistance in Cibitoke, Burundi"

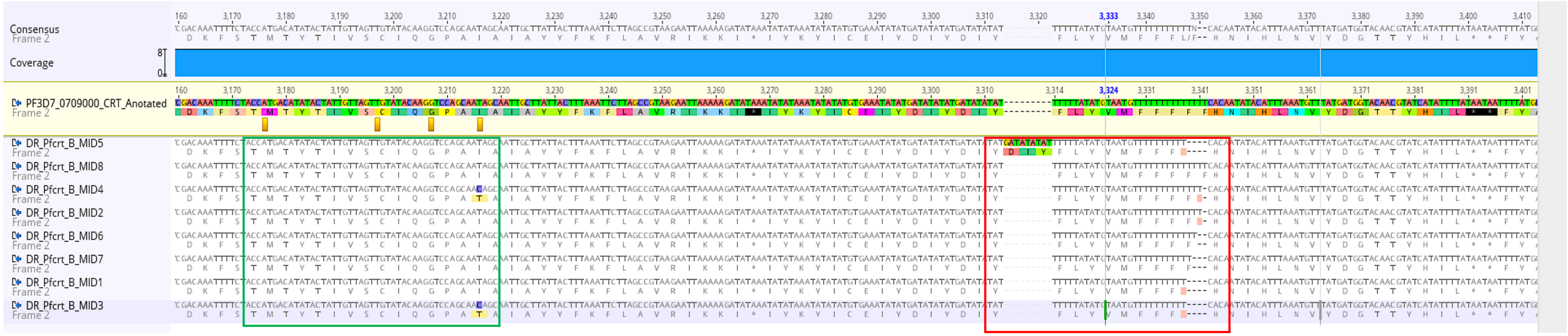


**Supplementary Figure S1.** Alignment of *pfcrt* amplicons spanning codons 343–356 (green box), used for SNP-based classification of wild-type or mutant alleles. Although several haplotypes (e.g., MID5, MID8, MID2, MID6, MID7, MID1) were wild-type at these codons, they exhibited distinct downstream indels (red box). Similarly, mutant haplotypes (MID4, MID3) also showed unique indels. While these insertions/deletions did not affect SNP-based allele classification, they contributed to overall haplotype diversity. Alignments visualized in Geneious Prime (v2025.0.3) using reference *pfcrt* (PF3D7_0709000).

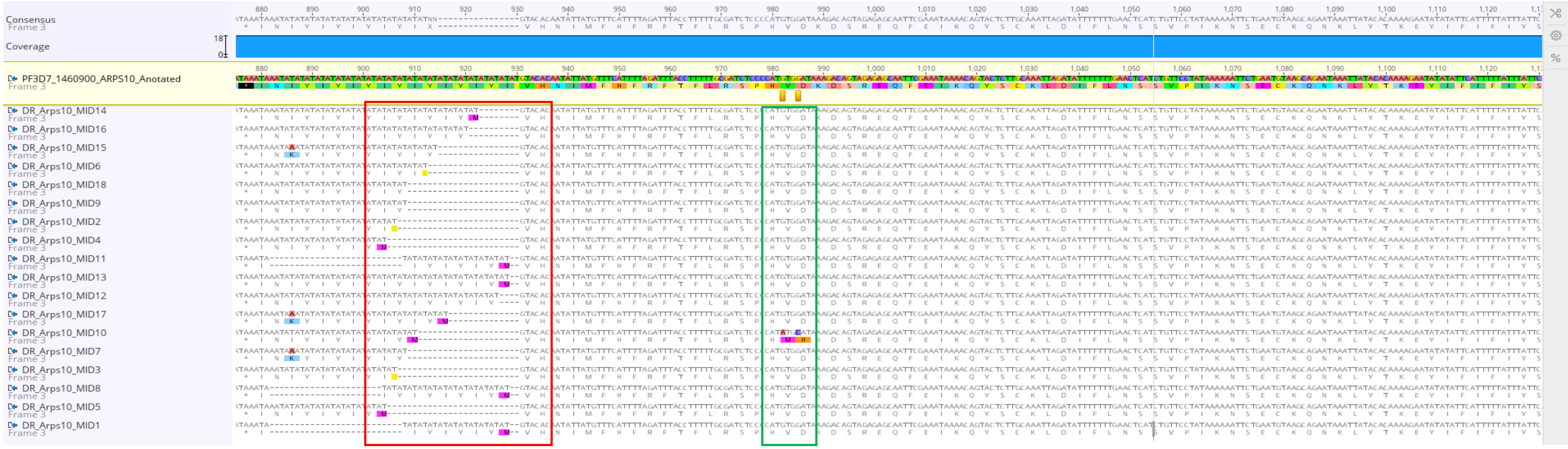


**Supplementary Figure S2**. Alignment of *pfarps10* amplicons spanning codons 127–128 (green box), used for SNP-based classification. All haplotypes except MID10 were wild-type at these positions but showed upstream insertions or deletions (red box). These indels did not affect SNP-based allele classification but contributed to haplotype diversity. Alignments were generated in Geneious Prime (v2025.0.3) using *pfarps10* reference PF3D7_1460900.

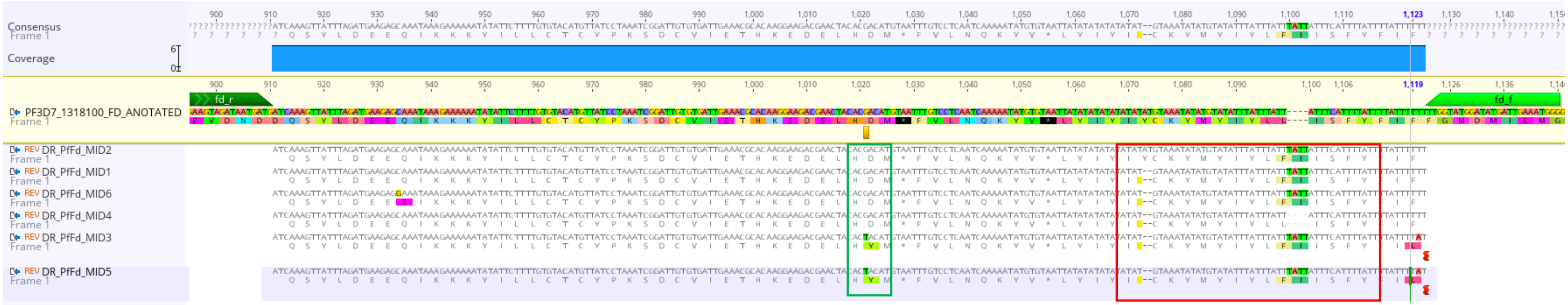


**Supplementary Figure S3.** Alignment of *pffd* amplicons highlighting codon 193 (green box), used for SNP-based classification. All haplotypes, except MID2, showed downstream deletions, and all but MID4 had insertions (red box). These indels did not affect allele classification at codon 193 but contributed to haplotype diversity. Alignments were visualized in Geneious Prime (v2025.0.3) using *pffd* reference PF3D7_1318100.

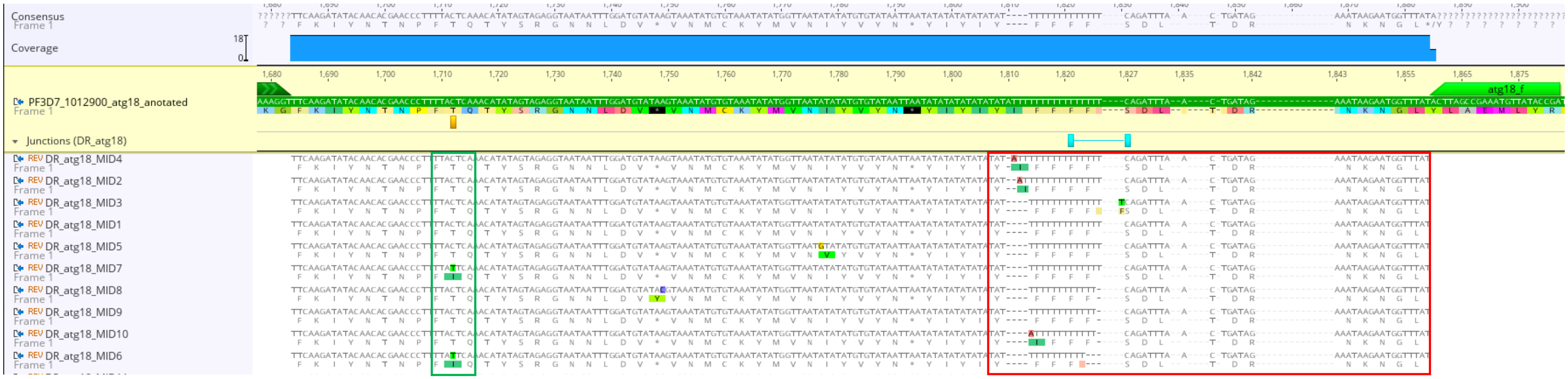


**Supplementary Figure S4.** Alignment of *pfatg18* amplicons highlighting codon 38 (green box), used for SNP-based classification. All haplotypes except MID6 and MID7 were wild-type at this position but exhibited downstream insertions and deletions (red box). These indels did not affect allele classification but added to haplotype diversity. Alignments were visualized in Geneious Prime (v2025.0.3) using *pfatg18* reference PF3D7_1012900.

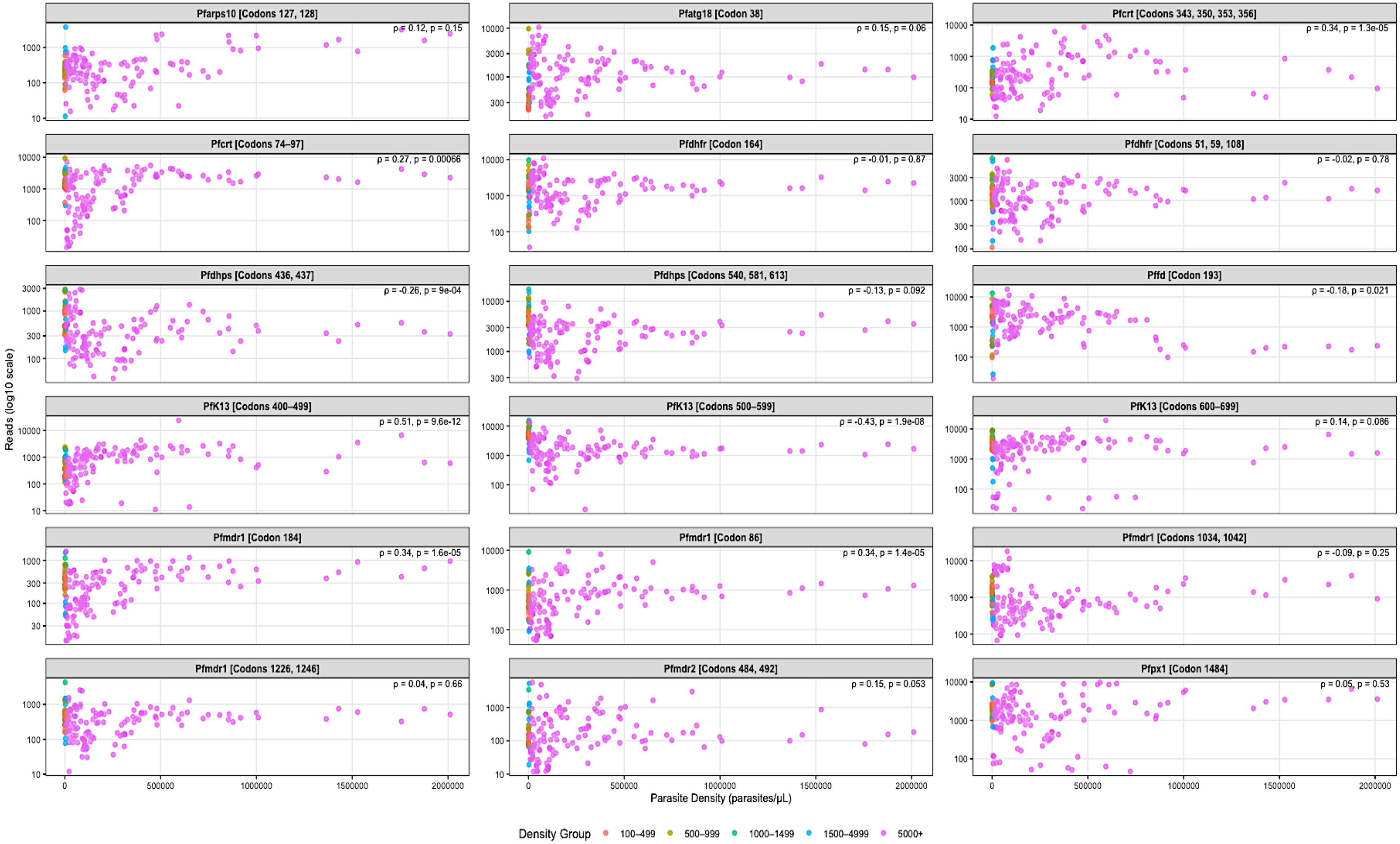


**Supplementary Figure S5.** Scatter plots showing marker-specific read counts plotted against parasite density (parasites/µL) across all samples passing sequencing QC (n = 157). Read counts are displayed on a log10 scale, and data are stratified by parasite density group and faceted by genotyping target. Spearman's correlation coefficient (ρ) and corresponding p-value are shown in each panel. Correlations between parasite density and read depth varied by marker. Moderate positive associations were observed for a subset of loci, including *PfK13* [Codons 400–499], *Pfcrt* [Codons 343–356], and *Pfmdr1* [Codons 86 and 184], whereas other targets (e.g., *Pfmdr1* [Codons 1034, 1042], *Pfdhps*, and *Pfdhfr*) showed weak, negligible, or negative associations. These patterns indicate locus-specific variability in amplification and sequencing performance rather than a consistent density-dependent effect on read depth. The plots serve as quality-control visualizations to assess marker performance and read yield across the observed range of parasite densities.

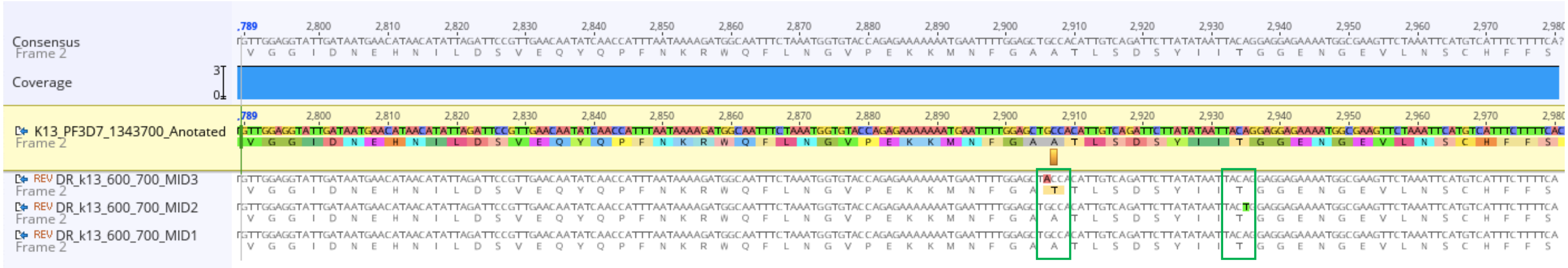


**Supplementary Figure S6.** Alignment of *pfkelch13* amplicons spanning codons 600–700. Nucleotide and amino acid alignment of *Pfkelch13* sequences showing a nonsynonymous A676T mutation (GCC→ACC) and a synonymous T685T mutation (ACA→ACT), highlighted in green boxes. A676T was observed in haplotype MID3, while MID1 and MID2 were wild-type at both positions. Alignments were visualized in Geneious Prime (v2025.0.3) using the *pfkelch13* reference (PF3D7_1343700).

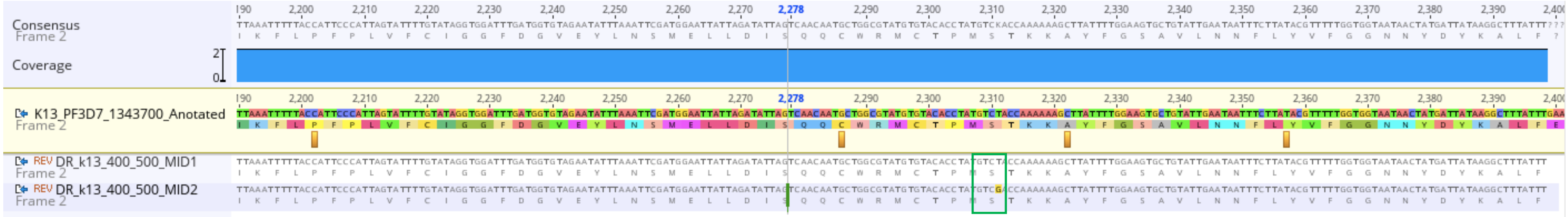


**Supplementary Figure S7.** Alignment of *Pfkelch13* amplicons spanning codons 400–500. Nucleotide and amino acid alignment of *pfkelch13* sequences showing a synonymous S477S mutation (TCT→TCG) in haplotype MID2. Alignments were generated in Geneious Prime (v2025.0.3) using the *pfkelch13* reference sequence (PF3D7_1343700).

A

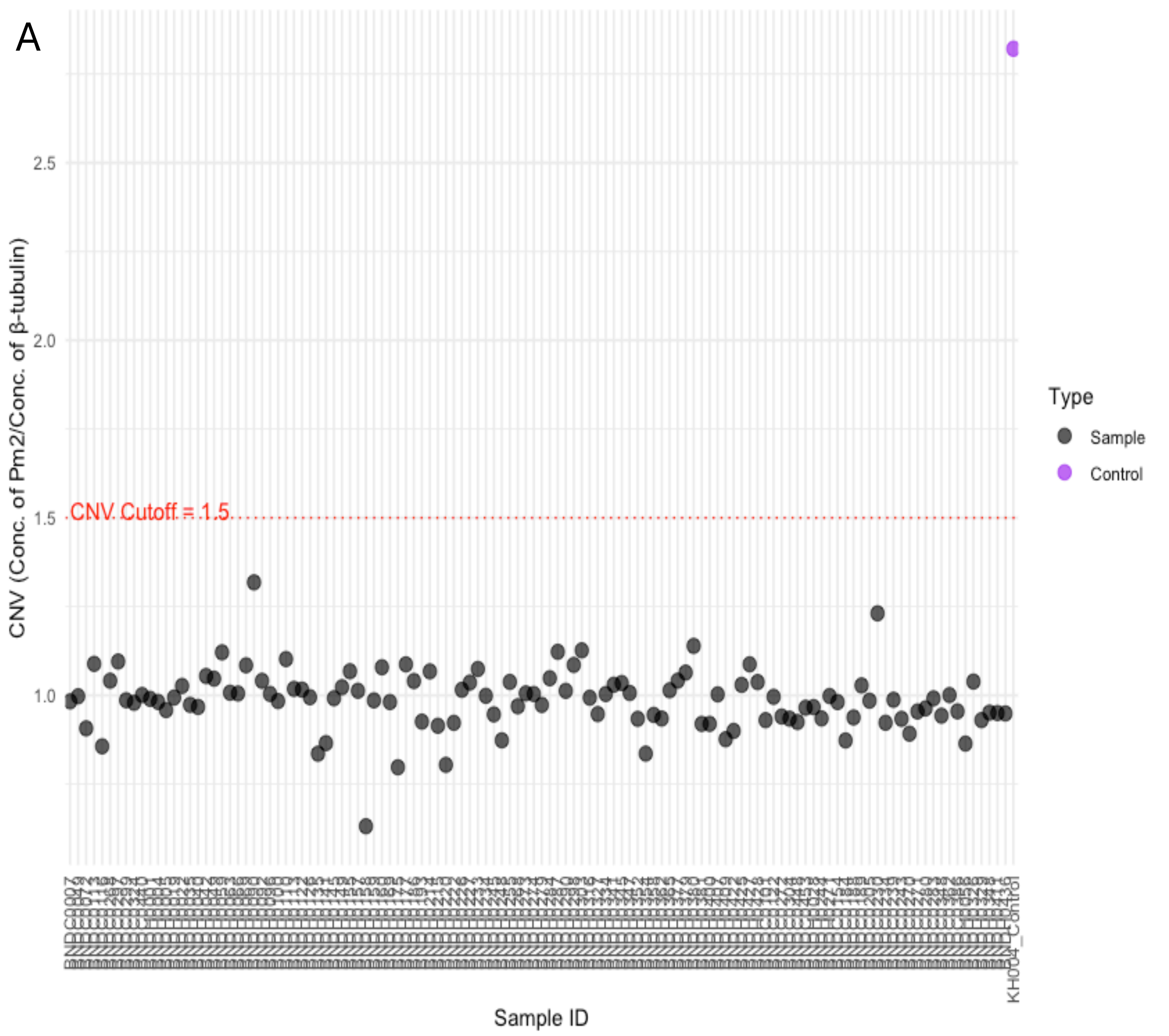
CNV (Conc. of Pm2/Conc. of β-tubulin)
2.5
2.0
1.5
1.0
CNV Cutoff = 1.5
Sample ID
KH004_Control
Type
Sample
Control

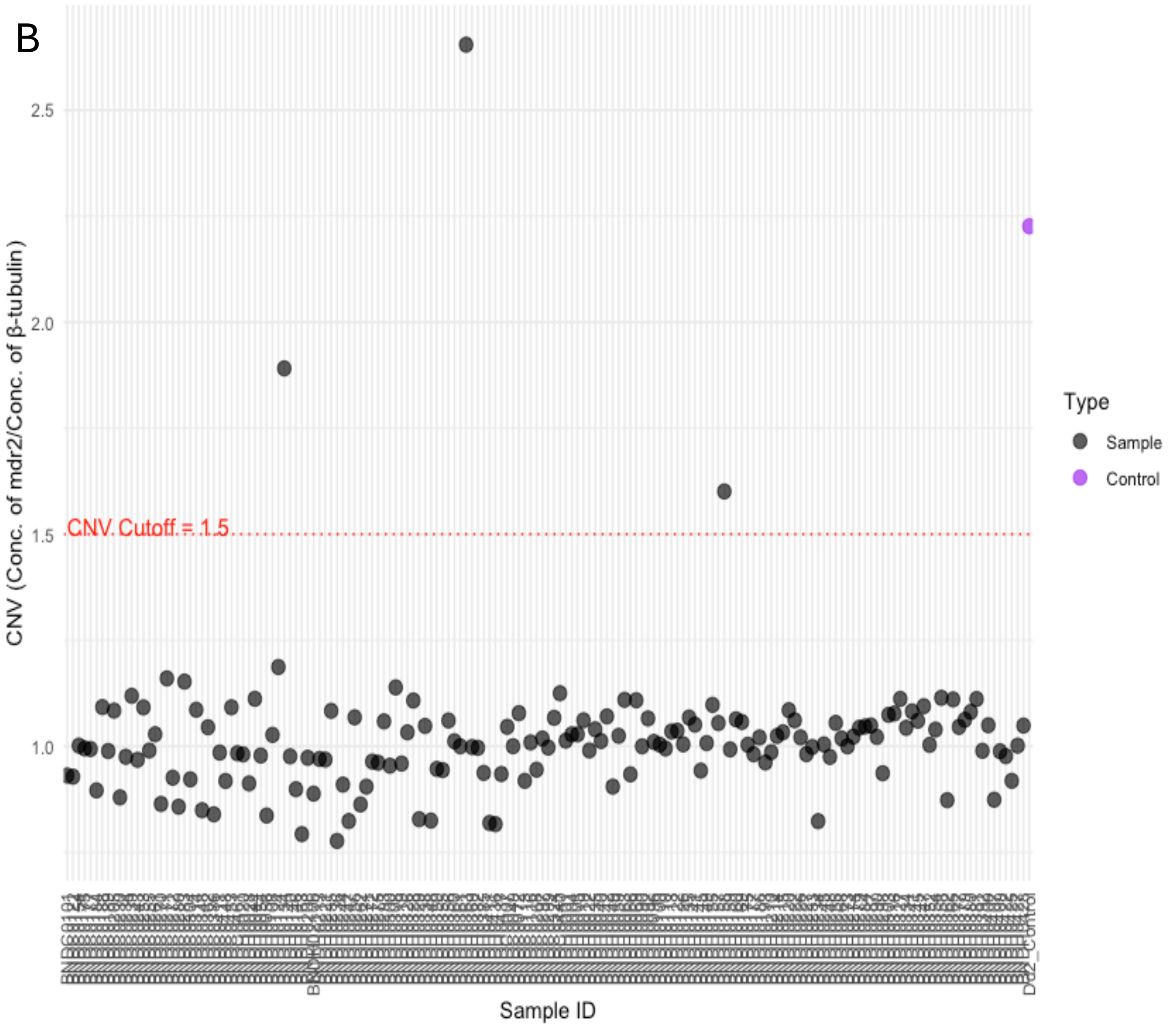


**Supplementary Figure S8. Copy number variations in (a) *plasmepsin 2* and (b) *Multidrug resistance gene 1*.** None of the 157 samples exhibited gene duplication for *pfpm2*, while three samples showed copy number variations (CNVs) exceeding 1.5 in *pfmdr1*. Purple dots in the *pfpm2* and *pfmdr1* assays represent positive controls (KH004 and Dd2).
